# Disability-Adjusted Life-Years (DALYs) for Breast Cancer and Risk Factors in 195 countries: Findings from Global Burden of Disease Study 2017

**DOI:** 10.1101/2020.04.02.20050534

**Authors:** Jieyu Liu, Jiaxiang Wang

## Abstract

**Background:** Breast cancer is a leading cancer burden for females. In order to picture the patterns and time trends of the breast cancer burden across different regions, the disability-adjusted life years (DALYs) were used to estimate breast cancer burden in 1990–2017.

**Methods:** Data from Global Burden of Disease Study 2017 (GBD 2017) was used to estimate the disability-adjusted life-years (DALYs) for the burden of breast cancer by locations, regions, years (with corresponding 95% uncertainty intervals [UI]). Besides, the associated potentially modifiable risk factors were estimated to provide targeted means for controlling the burden of breast cancer.

**Results:** All-age numbers of DALYs reached over 17.42 million years in 2017 globally, despite the decreasing trends in all-age and age-standardized rate. The rates for DALYs was the highest in Western Sub-Saharan Africa [694.23 (534.43 to 906.05)] in 2017. High fasting plasma glucose [1.07 million (0.20 to 2.43) DALYs] and high body-mass index (BMI) [0.81 million (0.27 to 1.53) DALYs] have become great attributors to DALYs of breast cancer in 2017.

**Conclusions:** The levels and trends in causes of DALYs of breast cancer, generally show similiarities between 2007 and 2017, although differences exist. The differences observed countries can be attributed high fasting plasma glucose and high body-mass index across the world. Concerned efforts at national and regional levels are required to tackle the emerging burden of breast cancer in the world.

## Introduction

As one of the most common cancer in women, breast cancer claiming 181,004 lives and resulting in 17.7 million disability-adjusted life years (DALYs), making it one of the most severe burdensome cancer globally. The incidence case of breast cancer in women will reach approximately 3.2 million by 2050[1]. Without doubt, breast cancer is a great threat to women’s health around the world[2].

Previous study from GBD 2016 showed that deaths from breast cancer increased from 1990 to 2016 and the incidence of breast cancer was higher in developed countries (high/very high HDI)[3]. However, this investigation only included incidence, deaths and mortality-to-incidence ratio (MIR), which was used as representative indicator of breast cancer survival, from available data and, in order to select incidence great than 1000 cases in 2016, they only recruited 102 countries for analysis[3]. However, the age-standardized incidence rates in some countries with less than 1000 incidence cases had increased drastically during the past periods[3].

Furthermore, Disability adjusted life years (DALYs), a summary measure of total health loss, was the sum of years of life-disability (YLDs) and years of life-lost (YLLs) for location, year, age and cause, which may be considered a better reflection of the burden of suffering caused by a disease than mortality rates[4]. An age-weighting function is used in DALY computation and values life years differently depending on the age of illness onset[5]. They are a useful analytical tool in cost-effectiveness analysis and are commonly used for comparison purposes[6]. Of note, the risk factors contributed for breast cancer globally and in different regions are disparated, which gain more practical application value for preventing breast cancer.

This paper aims to understand the burden of breast cancer in 195 countries, and the burden of breast cancer is evaluated by understanding the disability adjusted life year (DALY), which is calculated using years of life lost (YLL) and years lost due to disability (YLD), according to Global Burden of Disease Study (GBD) 2017. Furthermore, attributed risk factors for breast cancer varied significantly across different regions and time periods.

## Methods

### Data sources

Global Burden of Diseases, Injuries, and Risk Factors Study (GBD 2017) data were used to analysis DALYs for breast cancer. GBD uses all available up-to-date epidemiological data including a comprehensive assessment of incidence, prevalence, and years lived with disability (YLDs) and improved standardized methods to provide a comparative assessment of health loss across 359 diseases and injuries and 73 age-and-sex groups for 195 countries and territories[7].

As with GBD 2017, this analysis adheres to the Guidelines for Accurate and Transparent Health Estimates Reporting (GATHER) standards developed by WHO and others[8], which include recommendations on documentation of data sources, estimation methods, and statistical analysis. All data sources (except those protected by data usage agreement), code, and results are publicly available online. A comprehensive explanation of inclusion and exclusion criteria and limitations has been previously described elsewhere[9]. The Ethics Committee at the global

Council of Medical Research and the ethics committee of the Public Health Foundation approved the work of this initiative.

### Burden of breast cancer estimation

The GBD study uses the disability-adjusted life-years (DALYs), which is the sum of years of life lost due to premature mortality (YLLs) and years of life-disability (YLDs)[10], as a summary metric of population health loss, reflecting years of healthy life lost due to morbidity and mortality. Prevalence, deaths, and DALYs numbers were reported as absolute numbers and as age-standardized rates with 95% uncertainty intervals. Changes in absolute prevalence, deaths, and DALYs numbers are reported for 2007 and 2017 estimates. Details of the methods of the GBD study have been published elsewhere[11]. Using these data as primary inputs, DisMod-MR 2.1, a Bayesian meta-regression instrument, used a log rate and incidence-prevalence-mortality mathematical model to develop internally consistent epidemiological models. In view that empirical data were unequally distributed between age groups, and regions, DisMod-MR was able to impute age-specific estimates from available data, informed by expert prior information such as age of onset as well as data for comparable populations[7]. Statistical code used for GBD estimation was publicly available online.

### Geographical units and time periods

The GBD is hierarchically organized by geographic units or locations, with 7 super-regions, 21 regions nested within these super-regions, and 195 countries or territories within 21 regions. For the purposes of statistical analyses, we further grouped regions into seven super-regions (central Europe, eastern Europe, and central Asia; high income; Latin America and Caribbean; north Africa and Middle East; south Asia; southeast Asia, east Asia and Oceania; and sub-Saharan Africa)[12]. Subnational estimates for countries with populations larger than 200 million people (measured using our most recent year of published estimates) that have not yet been published elsewhere are presented wherever estimates are illustrated with maps but are not included in data tables.

### Statistical Analysis

Based on age-standardized rates on the GBD global reference population[13], both age-standardized rates and percentage change in age-standardized rates by DALYs were presented, since age-standardized rates allowed comparisons over time and between regions after adjusting for the differences in the age structure of the population. To estimate YLDs, GBD 2017 used a Bayesian metaregression method, DisMod-MR 2.1. It was designed to address statistical challenges in the estimation of nonfatal health outcomes, and for the synthesis of sparse and heterogeneous epidemiological data. The sequence of estimation occurs at five levels: global, super-region, region, country, and, where applicable, subnational locations. At each level of the cascade, the DisMod-MR 2.1 computational engine enforces consistency between all disease parameters. 95% uncertainty intervals (UIs) were calculated on the basis of 1000 draws from the posterior distribution of each step in the estimation process using the 2.5th and 97.5th percentiles of the ordered 1000 values.

## Results

### Age-standardized DALYs and Percentage change in age-standardized DALYs

Globally, DALYs of breast cancer had claimed 17.42 billion (95%UI 16.62 to 18.38) cases in 2017, with a 1.35% (95%UI -6.51 to 2.41) percentage reduction in age-standardized DALYs rate, per 100,000 population over the ten years. Of note, breast cancer related-DALYs contributed 1.78 billion (95%UI 1.64 to 1.94) in low SDI regions. While in high SDI regions increased to 4.24 billion (95%UI 4.03 to 4.46) (**Table 1**). The age-standardized DALYs rates were 694.23 in Western Sub-Saharan Africa and 692.47 in Oceania, per 100,000 DALYs. Whereas east Asia (257.54) and high-income Asia Pacific (288.94) remained the lowest in this regard (**Figure 1**).

**Table 1:**
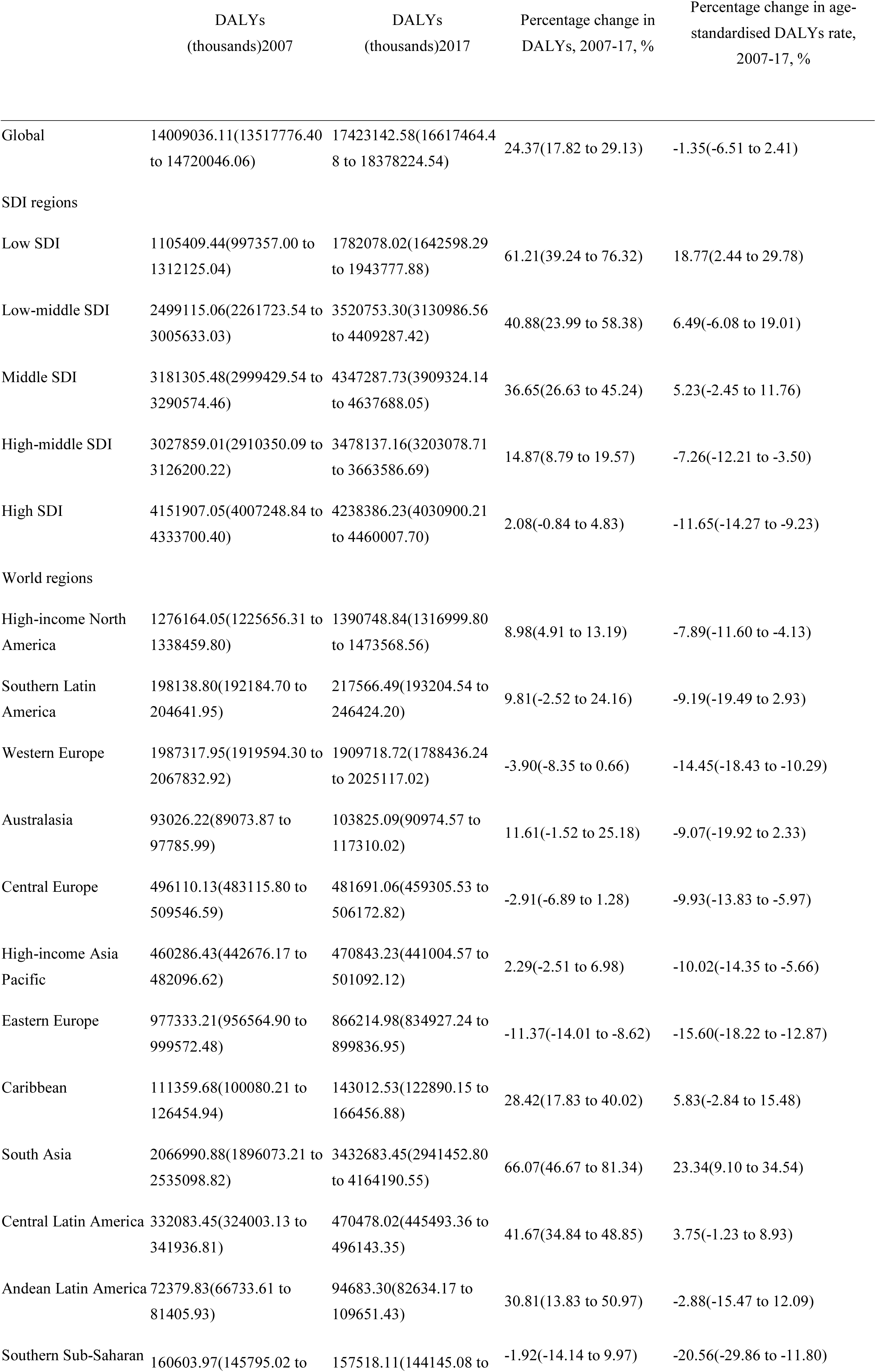
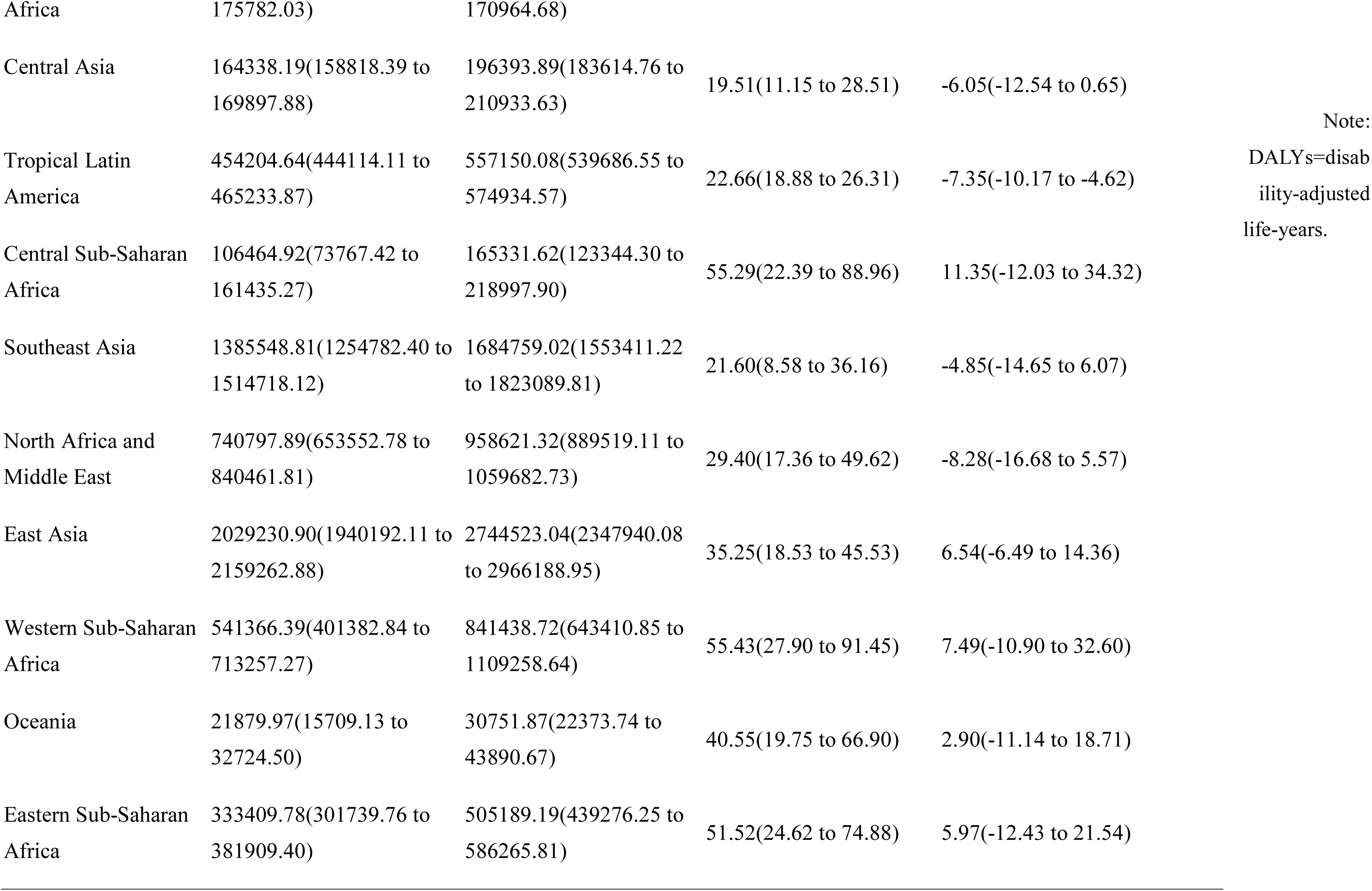
**DALYs per 100 000 population in 2007 and 2017, percentage change and percentage change in age-standardized rates for breast cancer during 2007–2017 by regions**.

**Figure 1:**
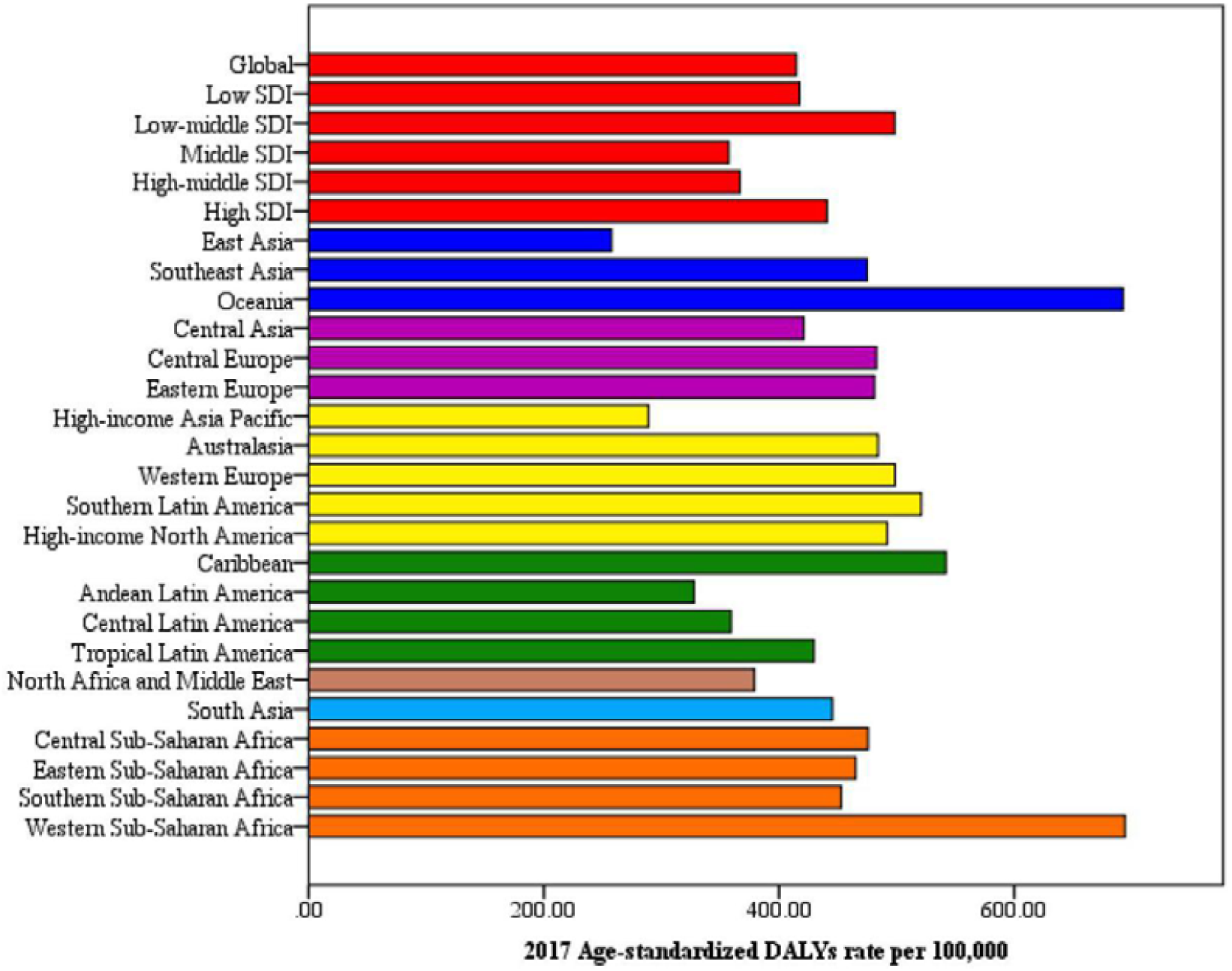
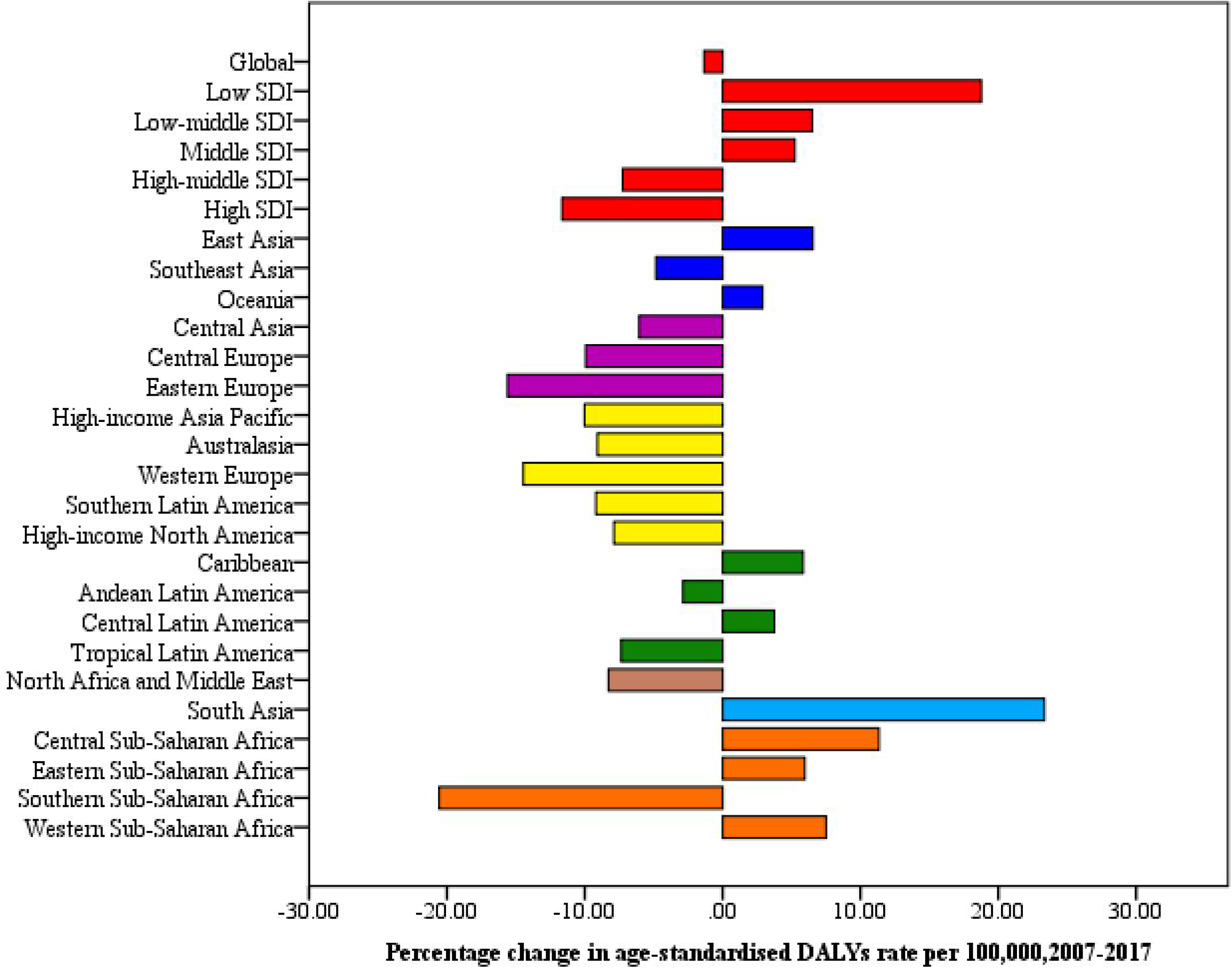
**Age-standardized DALYS rate in 2017 and percentage change in DALYS age-standardized rate by SDI and world regions, 2007–17. DALYs=disability-adjusted life-years**.

During 2007-2017, the percentage change in age-standardized DALYs rate was 18.77(2.44 to 29.78) in low SDI regions, while in high-middle and high SDI regions, rates of DALYs decreased by -7.26 (-12.21 to -3.50) and -11.65 (-14.27 to -9.23), respectively (**Table 1**). Besides, rates of DALYs in south Asia increased by 23% (9.0 to 35) per 100,000, whereas a decrease was observed in Southern Sub-Saharan Africa by 21% (-30 to -12) (**Figure 1**).

We noticed geographical variations in the highest and lowest DALYs (**Figure 2 and Supplementary Table 1**). Breast cancer burden was the highest in the Bahamas (1003.72), Pakistan (979.29), Tonga (925.72), and Nigeria (910.61) in 2017, as shown in **Figure 3**. During 2007-2017, the five leading countries with the largest increased fluctuations in terms of age-standardized DALYs rate were Grenada [+32.93% (16.65 to 50.09)], American Samoa [+22.89% (2.54 to 44.67)], and Jamaica [20.24% (-5.75 to 50.89)] (**Figure 3 and Supplementary Table 1**).

**Figure 2:**
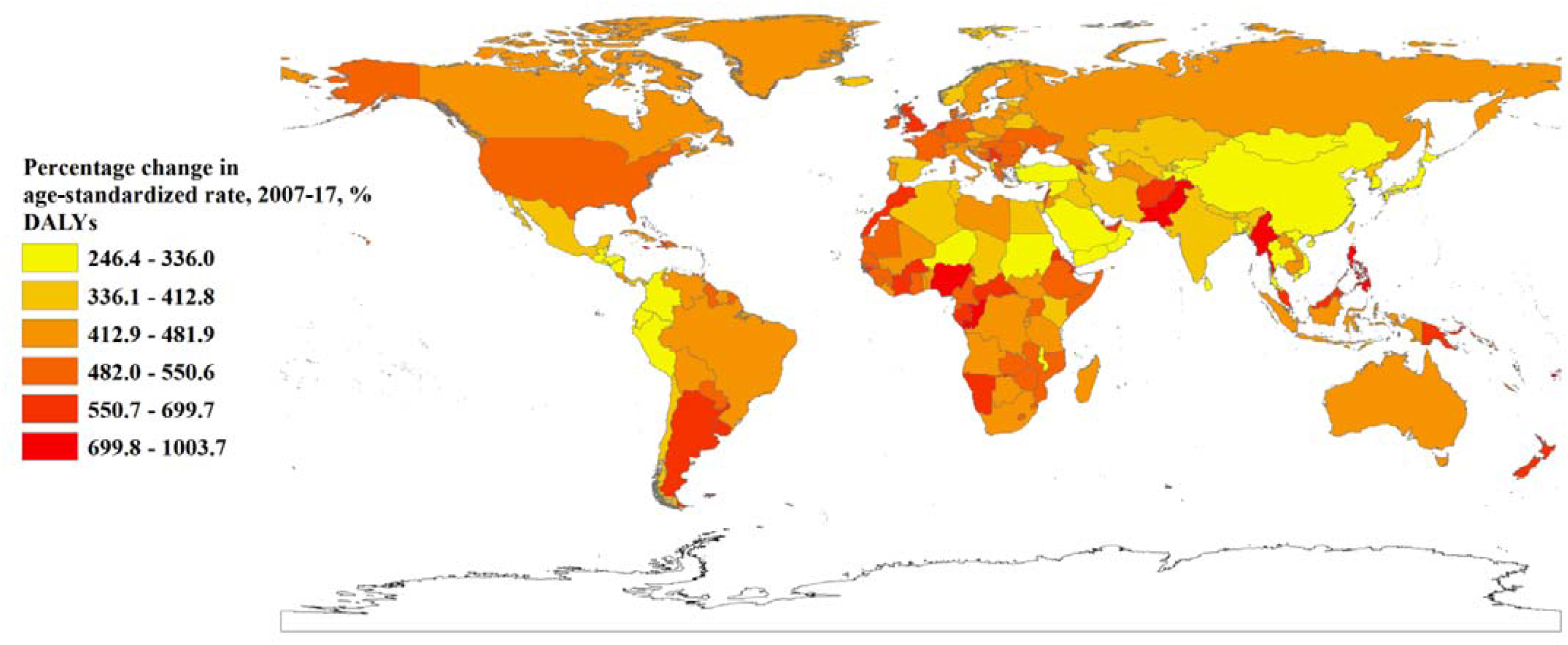
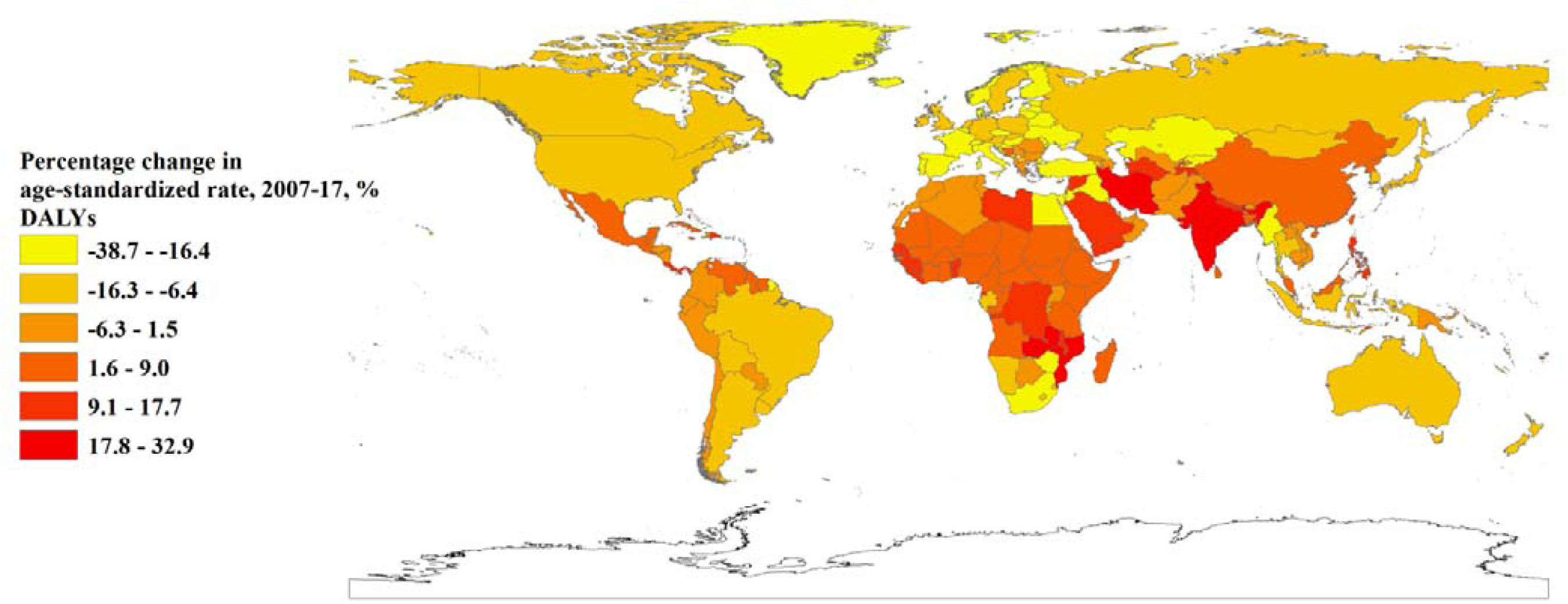
**Age-standardized rate in 2017 and percentage change in age-standardized rate by location, 2007–17. Age-standardized rate changes due to DALYs, Percentage change in age-standardised rate changes in DALYs. DALYs=disability-adjusted life-years. FSM=Federated States of Micronesia. Isl=Islands. LCA=Saint Lucia. TLS=Timor-Leste. TTO=Trinidad and Tobago. VCT=Saint Vincent and the Grenadines**.

**Figure 3:**
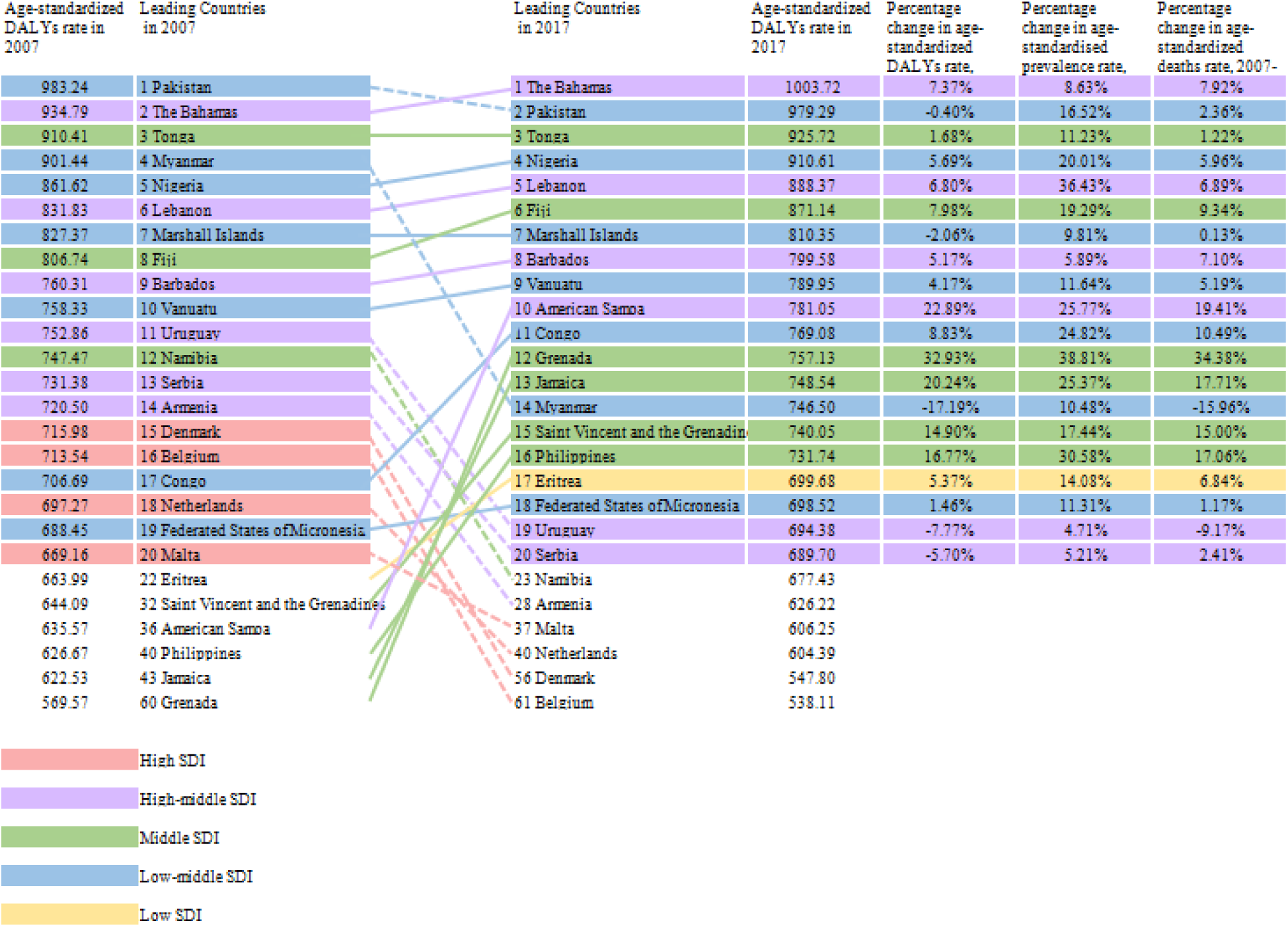
**The percentage change in age-standardized DALYs rate, 2007-17 of the top 20 countries with the highest age-standardized rate by DALYs in 2007 and 2017**.

Almost half the breast cancer-related DALYs occurred in people aged 50–54 years in low SDI countries. However, women aged 25-29 years in high SDI and high-middle SDI regions contributed greatly of all-age DALYs change (**Figure 4**).

**Figure 4:**
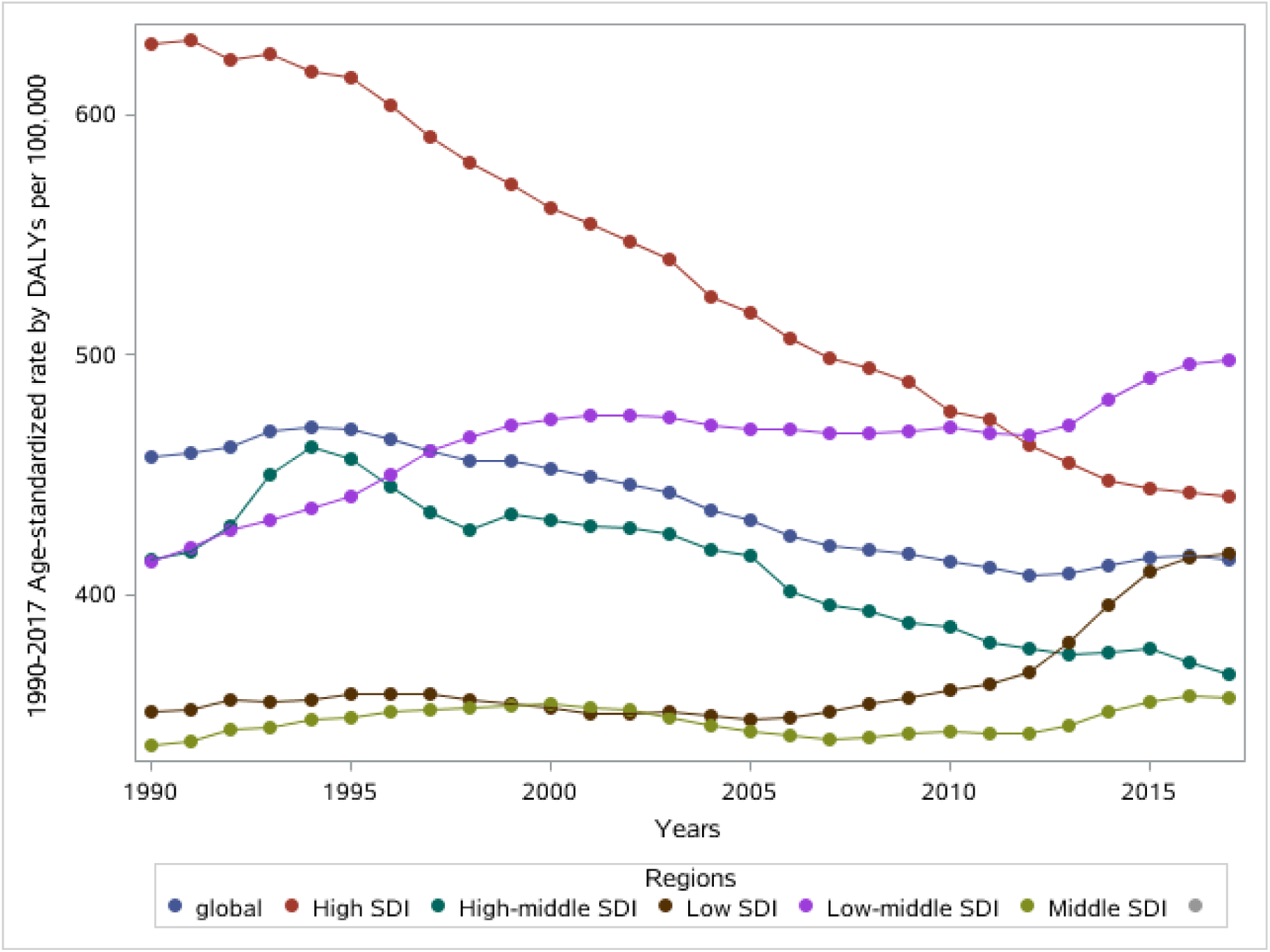
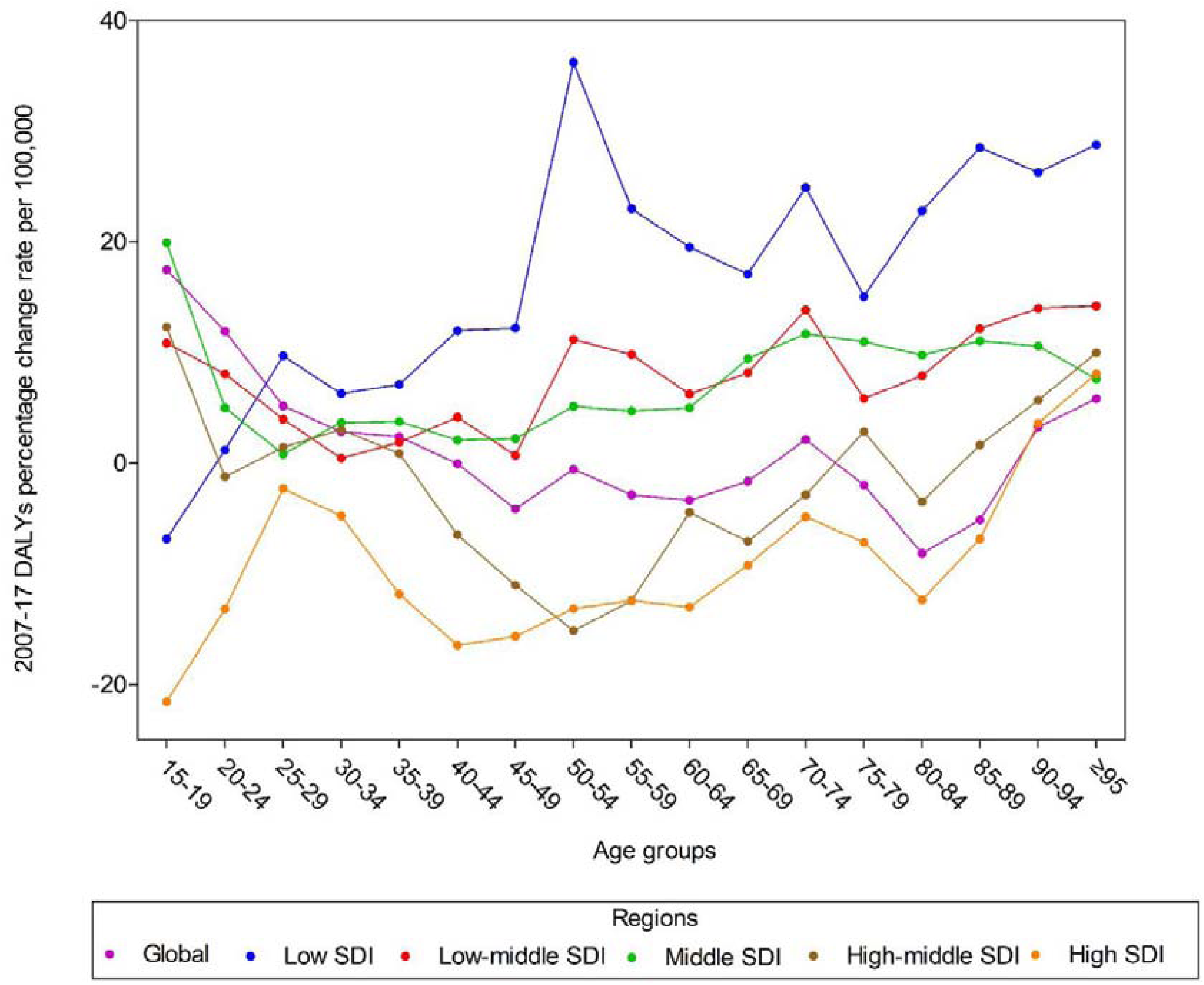
**Age-standardized rate by DALYs between 1990 and 2017 at the global level by SDI regions. Percentage change in DALYs rate at the global level by SDI regions, 2007–17 across different age groups between 15 to over 95 years old**.

### Risk factors for DALYs of breast cancer

Globally, high fasting plasma glucose and high BMI increased to be the leading causes of breast cancer during 2007-2017 (**Table 3 and Figure 5**). Of note, low physical activity contributed greater in low-income and low middle-income countries.

**Table 3:**
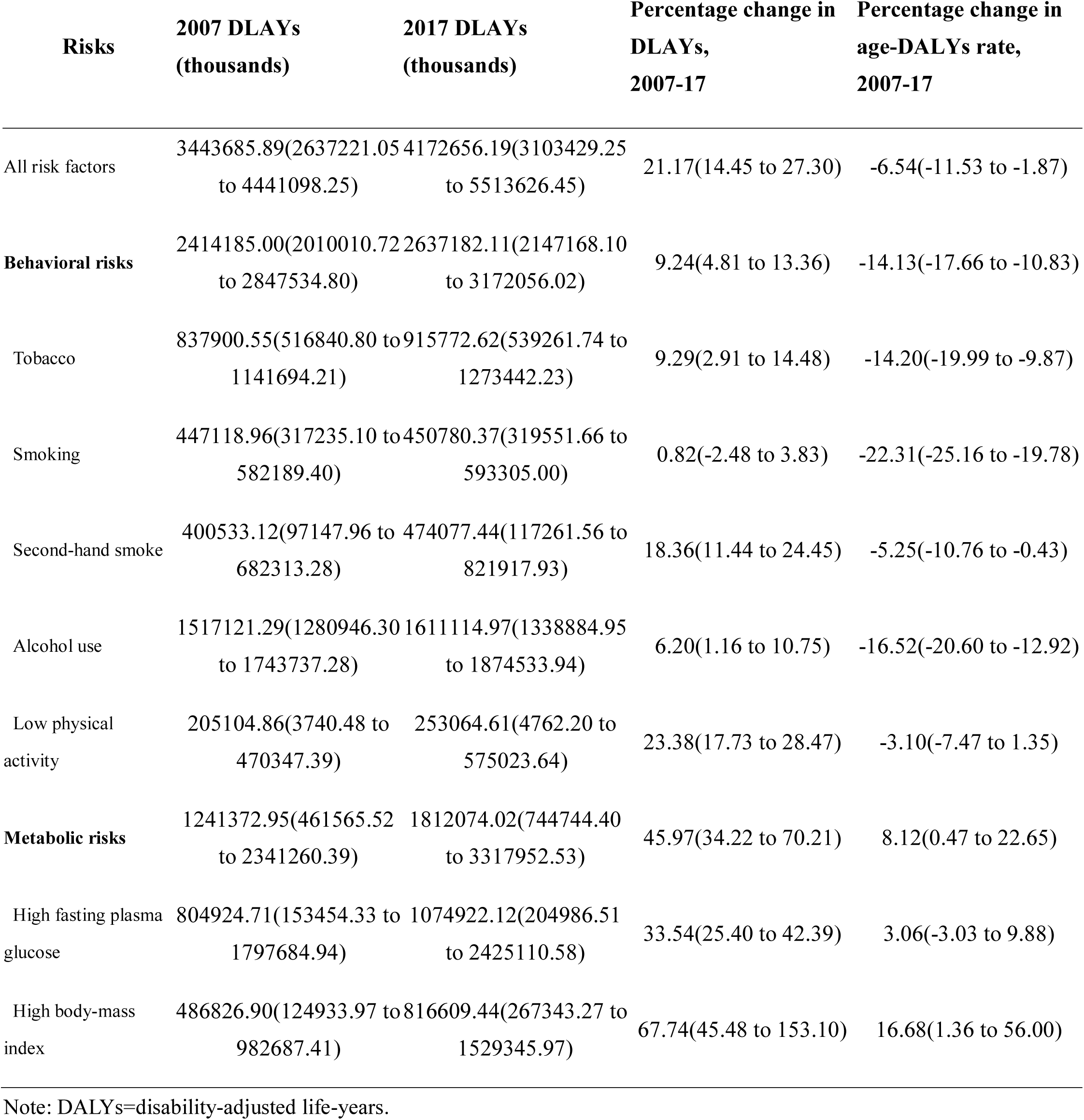
**Global all-age attributable DALYs, 2007**–**17, and percentage change of DALYs and age-standardised and DALY rates and death rates, 2007**–**17, for all risk-outcome pairs**.

**Figure 5:**
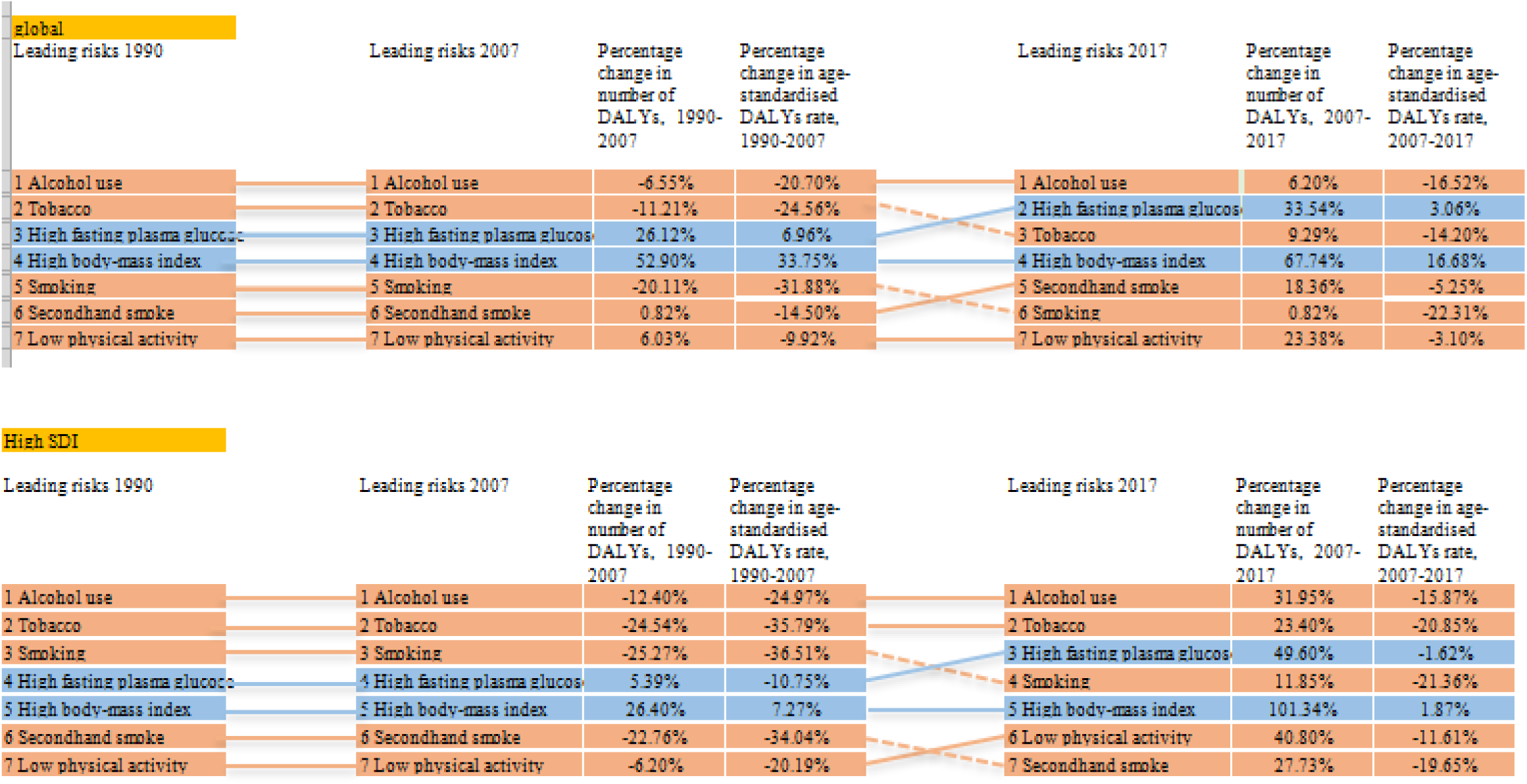
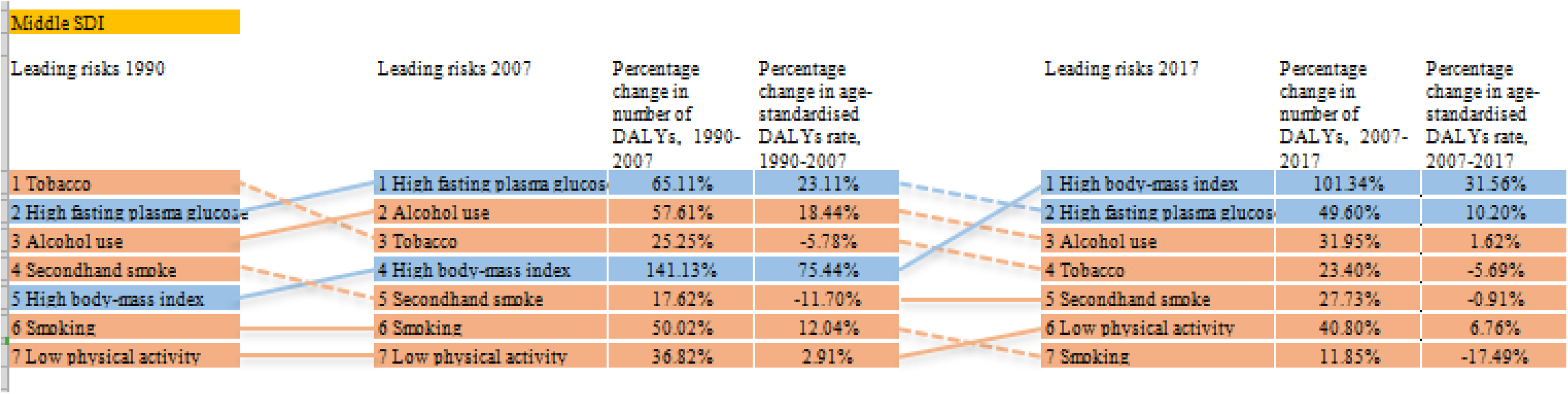
**Leading risk factors of breast cancer by attributable DALYs at the global level (A), in middle SDI region (B) and high SDI region (C) in 1990, 2007, and 2017 for females. Risks are connected by lines between time periods; solid lines are increases and dashed lines are decreases. Statistically significant increases or decreases are shown in bold (P<0·05). DALYs=disability-adjusted life-years**.

## Discussion

On a global scale, about 6.52 million female were diagnosed with breast cancer with 5.21% increment from 2007, among them 17.42 million DALYs cases (decreased 1.35% from 2007) in 2017. The trends for global breast cancers burdens differed among the SDI quintiles. Rising trends in DALYs has been consistently found in low SDI regions between 2007 and 2017, but declining trends for DALYs indicators were observed in high SDI regions. When considered about risk factors, high fasting plasma glucose and high BMI are concerned to become the great contributors of breast cancer by attributable DALYs around the world.

On the contrary, previous study indicated that 59% of breast cancer cases occurred in developed countries (north of America, Europe, Australia, New Zealand and Japan) in 1990[14], The prevalence rates were the highest in developed countries led by the Netherlands and Belgium in 2016[3]. In 2017, Netherlands still had the highest age-standardized prevalence rates. Though the prevalence rate was relatively high in developed countries, the age-standardized prevalence rate decreased significantly. In several developed countries, including the United States, Canada, the United Kingdom, France, and Australia, the fall in prevalence in 2000s was partly attributable to declines in the use of postmenopausal hormonal treatment after publication of the Women’s Health Initiative trial linking postmenopausal hormone use to increased breast cancer risk[15].

A worrying pattern was also presented in terms of different age, the odds of developing breast cancer were the highest in high SDI countries, this dramatic increase of breast cancer cases in young women is very important because the behavior of these tumors is in the majority of cases more aggressive in comparison with older women[16, 17]. Routine annual screening has been suggested to begin at 40 years of age, but only further research will establish whether this measure will be beneficial[18].

For breast cancer, the interplay of the risk factors to which the highest proportion of DALYs globally could be attributed, high fasting plasma glucose and high BMI. Elevated DALYs rate in high and middle SDI regions are both attributed to anthropometry (body fat distribution, and plasma glucose). These findings are generally consistent with previous study[19, 20], that obesity is associated with poor survival in of breast cancer[21]. We have found some slow progress in reducing tobacco consumption, but, no progress in lowering fasting plasma glucose and BMI, which has increased since 1990.

Our GBD study has several strengths. First, we employed data available from the Global Burden of Disease (GBD) study, which draws strength from data availability from cancer registries. Second, compared with the GBD 2016 of breast cancer[3], We have made substantial improvements in the estimation of life expectancy in GBD 2017 across our publication, including an independent estimation of population, a comprehensive update on fertility, adding a substantial amount of new data (from censuses, Demographic Surveillance Sites, and other sources), improvements to the GBD model life-table system, and enhancements to the modelling framework[22]. We calculated DALYs in the part of GBD 2017, which reflected years of healthy life lost for cancer better than the deaths rates[23]. Besides, we conducted the GBD study based on 195 countries within 21 regions to expand demographic units for the global burden of breast cancer. What’s more, only a good understanding of the distribution of risk factors can we better formulate the corresponding preventive measures for the specific countries.

Our GBD study has several limitations. For this study, inclusion of these and other risk factors might have resulted in an even greater proportion of breast cancer burden attributable to risk factors, because the missing factors are known to have a role in breast cancer. To address the knowledge gap in GBD, the next step would be to include previous health conditions by age, and cancer subtypes in analyses, in addition to at least some of the broader causes. We also aim to update such crucial evidence for breast cancer prevention at all levels every 2 years, including geographical regions, time periods, and further risk factors.

Despite these acknowledged and important data limitations, our study is crucial to quantify the global burden of breast cancer in 195 countries. We believe that our study will be of particular interest to health care program. As high fasting plasma glucose, high BMI, and low physical activity are important part of breast cancer’s etiology, it may be particularly suited to population-based interventions in order to improve the public self-discipline. In addition, future research needs to investigate the drivers of the observed heterogeneity in the epidemiological and demographic transitions.

## Data Availability

All data are available online

